# Standardized clinical data capture to describe cerebral palsy

**DOI:** 10.1101/2024.08.09.24311474

**Authors:** Susie Kim, Kelsey Steffen, Lauren Gottschalk-Henneberry, Jennifer Miros, Katie Leger, Amy Robichaux-Viehoever, Karen Taca, Bhooma Aravamuthan

## Abstract

**Objective:** To describe a standardized methodology for capturing clinically valuable information on young people with cerebral palsy (CP) from caregivers and clinicians during routine clinical care.

**Methods:** We developed a caregiver-facing intake form and clinician-facing standardized note template and integrated both into routine clinical care at a tertiary care CP center (https://bit.ly/CP-Intake-Methodology). We extracted this caregiver and clinician-entered data on people with an ICD10 diagnosis of CP seen between 3/22/23 and 12/28/23. We used this data to describe how CP manifests in this group and which medical features affected the odds of walking, oral feeding, and speech by age 5.

**Results:** Of 686 visits, 663 (97%) had caregiver- and clinician-entered data and 633 had a clinician-confirmed CP diagnosis (mean age 9.1, 53.4% Male, 78.5% White). It was common to have quadriplegia (288/613, 47.0%), both spasticity and dystonia (257/632, 40.7%), walk independently (368/633, 58.1%), eat all food and drink safely by mouth (288/578, 55.9%), and produce understandable speech (249/584, 42.6%). Cortical grey matter injury and duration of initial critical care unit stay affected the odds of walking, oral feeding, and speech (binary logistic regression, p<0.001).

**Conclusions:** We comprehensively captured caregiver and clinician-entered data on 97% of people seen in a tertiary care CP Center and used this data to determine medical features affecting the odds of three functional outcomes. By sharing our methodology, we aim to facilitate replication of this dataset at other sites and grow our understanding of how CP manifests in the US.

**Article summary:** Using caregiver and clinician-entered data on people seen in a tertiary-care CP center, we determined medical features affecting the odds of three functional outcomes.

**What’s known on this subject:** Detailed CP characterization can be limited if using population-based registries and retrospective chart review alone, including limited data on recently validated functional classification systems for CP.

**What this study adds:** We comprehensively captured caregiver and clinician-entered data on 97% of people seen in our CP Center to describe how CP manifests and show that cortical injury and initial ICU stay duration affect the odds of walking, oral feeding, and speech.

**Contributors Statement:** Susie Kim helped design the study, aggregated data, carried out data analyses, and critically reviewed and revised the manuscript.

Kelsey Steffen helped conceptualize and design the study and critically reviewed and revised the manuscript.

Lauren Gottschalk, Jennifer Miros, Katie Leger, Amy Viehoever, and Karen Taca helped design the study and critically reviewed and revised the manuscript.

Bhooma Aravamuthan conceptualized and designed the study, supervised data collection and analysis, drafted the initial manuscript, and critically reviewed and revised the manuscript.

## Introduction

Cerebral palsy is the most common childhood motor disability, affecting 2 of every 1000 children in the US.^1^ Given its high prevalence, it is critical to have a detailed understanding of how CP manifests. The Centers for Disease Control and Prevention last assessed CP prevalence in 2016^1^ using retrospective medical record review in targeted geographical regions. This methodology is valuable for collecting broad prevalence data but precludes detailed CP characterization due to variability in clinical documentation practices and the required personnel hours. The Cerebral Palsy Research Network (CPRN) facilitates multi-institutional prospective data entry as a part of clinical workflow,^2^ and currently partners with 35 tertiary care centers (34 in the US)^3^ to document at least five “essential” items required for all CP care.^4^ This helps identify large subpopulations of people with CP for further detailed analysis but may not provide detailed CP characterization in isolation. National CP registries outside the US have conducted detailed population-based prospective data collection, but typically require significant financial, personnel, and time investments.^5^ Data on more recently validated functional classification systems for CP have yet to be broadly collected in these registries and more specific data like medication history for tone and seizure management are also not typically captured.^5^ Therefore, acknowledging the strengths and gaps of the above approaches, there remains significant value in comprehensive and standardized data capture at large single centers caring for people with CP.

Here, we describe our methodology for standardized clinical documentation and prospective data collection in a pediatric tertiary care CP center. We use this data to see which medical factors affect the odds of three functional outcomes: walking, oral feeding, and speech. Our goals are to facilitate recapitulation of our methodology at other sites and contribute to detailed characterization of CP in the US.

## Methods

### Standard Protocol Approvals, Registrations, and Patient Consents

Human Subjects Research exemption was granted by the Washington University Institutional Review Board (ID#202309003, 09/11/2023).

### Creation of the caregiver-facing intake form and clinician standardized note template

Data was collected in the Cerebral Palsy and Mobility Center affiliated with St. Louis Children’s Hospital and the Washington University Department of Neurology. The CP Center is staffed by 2 pediatric movement disorders neurologists and a pediatric nurse practitioner with multiple years’ experience caring exclusively for people with CP and movement disorders.

The caregiver-facing intake form and clinician note template were designed to include medical issues valuable for CP care as enumerated by the American Academy of Pediatrics Council on Children with Disabilities.^6^

Clinician documentation, exclusively in Epic (Epic Systems Corporation), included:

- Diagnosis (CP or not)
- Predominant and other tone/movements (determined using published descriptions^7–13^ and the Hypertonia Assessment Tool^14^)
- Anatomic distribution (e.g. diplegia, quadriplegia)
- Etiology/-ies
- Functional ability using tools validated for CP:

`o` Gross Motor Functional Classification System (GMFCS, valid for all ages)^15,16^
`o` Manual Ability Classification System (MACS, age 4+)^17^
`o` Communication Function Classification System (CFCS, age 2+)^18^
`o` Motor verbal speech assessed using the Viking Speech Scale (VSS, age 4+)^19^
`o` Visual Function Classification System (VFCS, age 1+)^20^
`o` Eating and Drinking Ability Classification System (EDACS, age 3+)^21^
- Brain MRI findings (for those with images in the EHR) using the MRI Classification System^22^ validated for CP.

Caregiver documentation included:

- Demographics (National Institutes of Health Common Data Elements^23^)
- Birth history
- Medication and botulinum toxin history for motor/tone management
- Orthotics and equipment
- Therapies
- Medical specialties seen and surgical history
- Seizure history
- Sleep, pain, mood, sensory, and social concerns
- Family history

To avoid diagnostic variability,^24^ clinicians agreed *a priori* to diagnose CP per the 2006 definition^25^ in any child with a disturbance to the developing brain sustained before 2 years of age who has or is predicted to have a resultant permanent and non-progressive motor disability.^26^ Clinicians also agreed to include predominant or pure hypotonia as CP phenotypes, in line with half of CP registries globally.^5,24^ Area Deprivation Index (ADI) was determined using 9-digit zip codes in the EHR.^27^

The intake form and note template (https://bit.ly/CP-Intake-Methodology) were iteratively revised by all members of the CP Center (including physical and occupational therapists, orthotists, seating and equipment specialists, a social worker, a nurse coordinator, and a certified nurse assistant) between 12/1/2022 and 3/1/23. Between 1/30/23 and 3/22/23, families seen in the CP Center piloted the form and provided their feedback during clinic visits. The intake form and note template were launched as routine clinical workflow on 3/22/2023. Clinician data entry for the MACS, CFCS, VSS, VFCS, and EDACS was additionally launched on 4/15/2023.

### Data capture

Inclusion criterion: People seen in the CP Center between 3/22/23 and 12/28/23 with a primary visit ICD10 diagnosis of CP (G80). Exclusion criteria: 1) No clinician-entered Smartlist data; 2) No caregiver-entered intake form data; and 3) Person does not have CP (via clinician-entered Smartlist designation).

Families are sent the intake form via a REDCap link (https://redcap.link/CP-intake-example) provided in a mailed flyer, MyChart message, email, phone call, and secure tablet device in the clinic waiting room. Completed intake form data is exported as a CSV file, converted into a readable format using a mail merge template in Microsoft Word, and then entered in the EHR by a certified nurse assistant (SK). Clinicians can pull this information into their Epic Smartphrase note template and then enter data in the note using Epic Smartlists. Smartlist selections are mapped to Smart Data Elements which populate an Epic Clarity Table for data export (https://bit.ly/CP-Intake-Methodology).

### Data analysis

Both clinicians and caregivers were queried about functional abilities, but only clinician-entered functional assessments were used for analysis. For some data elements, caregivers were asked to select all choices that apply, precluding differentiation between missing data and situations where no choices applied (denoted in the Tables as "none indicated”). Missing data percentages were otherwise calculated for each data element.

Data were analyzed descriptively (percentages and means with 95% confidence intervals, CIs) in Microsoft Excel. Binary logistic regression (SPSS, IBM, Armonk, NY) was used to assess whether five variables which can be determined in infancy (ADI, gestational age, birth weight, MRICS brain injury pattern, and etiology) affected the odds of: a) independent ambulation (GMFCS levels I-III vs. levels IV-V), b) ability to take nutrition orally (directly queried), and c) ability to produce verbal speech understandable to unfamiliar listeners (VSS levels I-II vs. levels III-IV).

To ensure that outcome variables were stable when assessed, only data collected for children 5 years and older were used for logistic regression analysis. Chi-square and Wald tests were used to determine significance of the model and model terms, respectively (p<0.05), and the Hosmer-Lemeshow test was used to estimate goodness of model fit (p>0.05). Adjusted odds ratios (ORs) and 95% CIs were calculated.

<u>Data sharing:</u> De-identified data will be shared with qualified investigators upon request. See https://bit.ly/CP-Intake-Methodology for all materials necessary to replicate our methodology.

## Results

Of 686 visits meeting inclusion criteria, 663 (97%) had caregiver- and clinician-entered data. Of these, 30 did not have CP, yielding data on 633 people with CP. The median percentage of missing data across all data elements was 1.9% (range 0-10.4%). Noting that clinicians began routinely entering MACS, CFCS, VSS, VFCS, and EDACS data in the clinical note 24 days into the 281-day data collection period, there is greater percentage of missing data for these functional classification systems (median 9.5%) than for other data (median 0.2%). As an indicator that caregivers completed the intake form fully, the last page of the intake form had a median missing data percentage of 0.8% (range 0.3-3.0%).

### Demographics

People with CP were 9.1 years old on average (95% CI 8.3 to 9.9 years old, range 0.5-23.2 years) and 53.6% (337/629) male. Most caregivers (523/629, 83.1%) identified their race and the race of the young person they cared for as White (492/627, 78.5%). A minority of caregivers had a bachelor’s degree or higher (251/614, 39.7%). Noting that higher ADIs indicate greater socioeconomic disadvantages, 233/633 (36.8%) had an ADI greater than the 75%ile for the country and 456/633 (72.0%) had an ADI greater than the 50%ile. Families lived an average of 68.1 miles from the CP Center (95% CI 58.5 to 77.8 miles) (Figure 1, Table 1).

**Figure 1.**
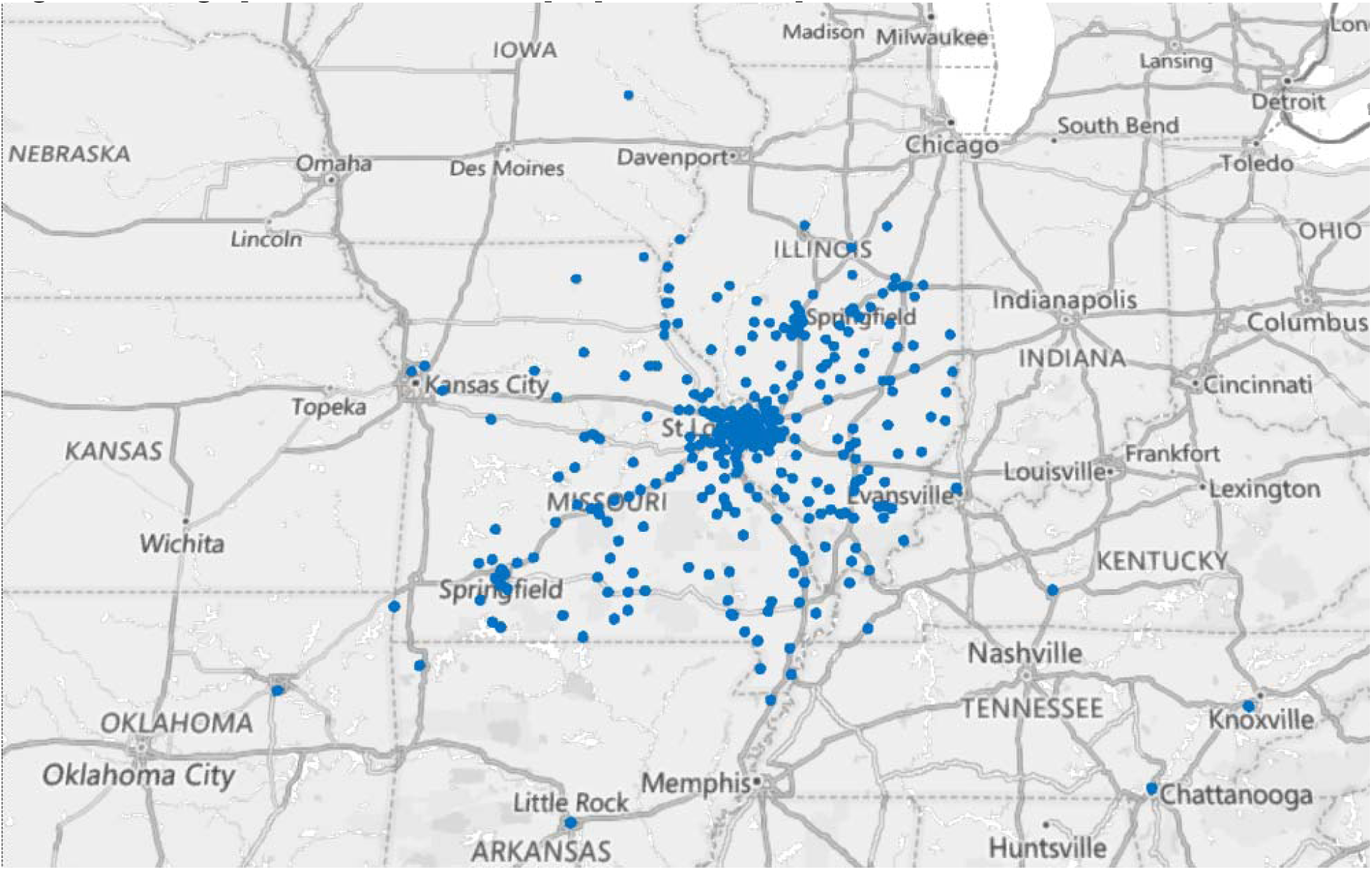
Geographical distribution of people with CP represented in this data set.

**Table 1.**
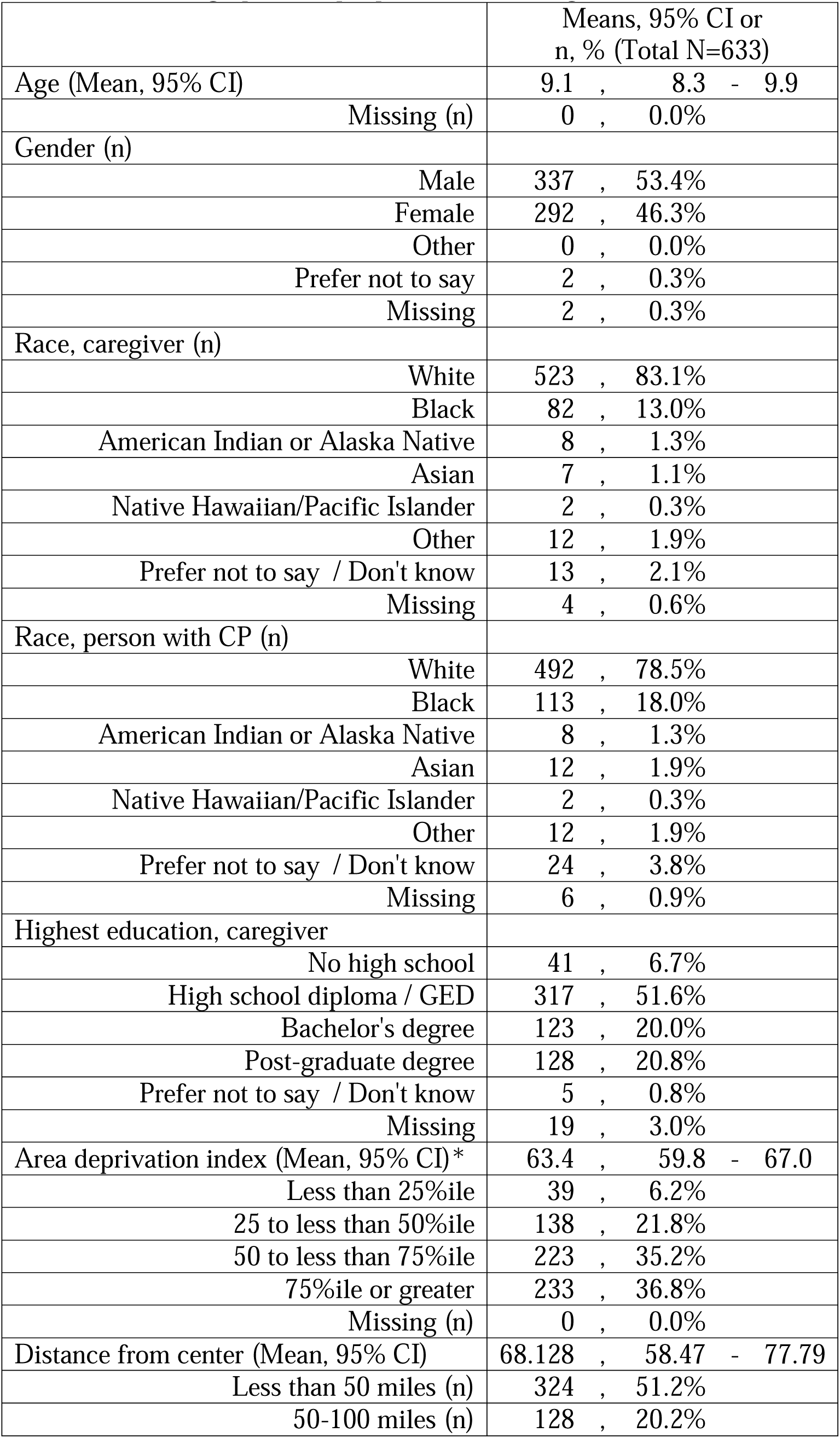
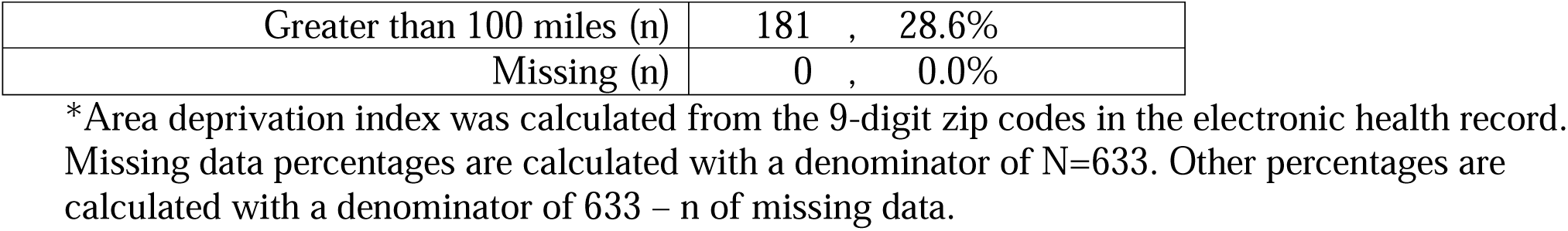
Demographics of people with CP (caregiver-entered)

|  | Means, 95% CI or<br>n, % (Total N=633) |
| --- | --- |
| Age (Mean, 95% CI) | 9.1 , 8.3 - 9.9 |
| Missing (n) | 0 , 0.0% |
| Gender (n) |  |
| Male | 337 , 53.4% |
| Female | 292 , 46.3% |
| Other | 0 , 0.0% |
| Prefer not to say | 2 , 0.3% |
| Missing | 2 , 0.3% |
| Race, caregiver (n) |  |
| White | 523 , 83.1% |
| Black | 82 , 13.0% |
| American Indian or Alaska Native | 8 , 1.3% |
| Asian | 7 , 1.1% |
| Native Hawaiian/Pacific Islander | 2 , 0.3% |
| Other | 12 , 1.9% |
| Prefer not to say / Don't know | 13 , 2.1% |
| Missing | 4 , 0.6% |
| Race, person with CP (n) |  |
| White | 492 , 78.5% |
| Black | 113 , 18.0% |
| American Indian or Alaska Native | 8 , 1.3% |
| Asian | 12 , 1.9% |
| Native Hawaiian/Pacific Islander | 2 , 0.3% |
| Other | 12 , 1.9% |
| Prefer not to say / Don't know | 24 , 3.8% |
| Missing | 6 , 0.9% |
| Highest education, caregiver |  |
| No high school | 41 , 6.7% |
| High school diploma / GED | 317 , 51.6% |
| Bachelor's degree | 123 , 20.0% |
| Post-graduate degree | 128 , 20.8% |
| Prefer not to say / Don't know | 5 , 0.8% |
| Missing | 19 , 3.0% |
| Area deprivation index (Mean, 95% CI)* | 63.4 , 59.8 - 67.0 |
| Less than 25%ile | 39 , 6.2% |
| 25 to less than 50%ile | 138 , 21.8% |
| 50 to less than 75%ile | 223 , 35.2% |
| 75%ile or greater | 233 , 36.8% |
| Missing (n) | 0 , 0.0% |
| Distance from center (Mean, 95% CI) | 68.128 , 58.47 - 77.79 |
| Less than 50 miles (n) | 324 , 51.2% |
| 50-100 miles (n) | 128 , 20.2% |
| Greater than 100 miles (n) | 181 , 28.6% |
| Missing (n) | 0 , 0.0% |

\*Area deprivation index was calculated from the 9-digit zip codes in the electronic health record. Missing data percentages are calculated with a denominator of N=633. Other percentages are calculated with a denominator of 633 – n of missing data.

### Motor tone types, distribution, and functional abilities

The most common tone types were spasticity (533/632, 84.3%), dystonia (369/632, 58.4%), and hypotonia (320/633, 50.6%) with 120/632 (19.0%) having hypotonia as their predominant tone type. Most people had a mixed tone/movement pattern (462/632, 73.1%) even when excluding hypotonia (348/584, 59.6%). The most common presentation was spasticity predominant tone accompanied by dystonia (257/632, 40.7%) (Table 2). Quadriplegia was the most common CP distribution (288/613, 47.0%) followed by diplegia (121/613, 19.7%) (Table 3).

**Table 2.**
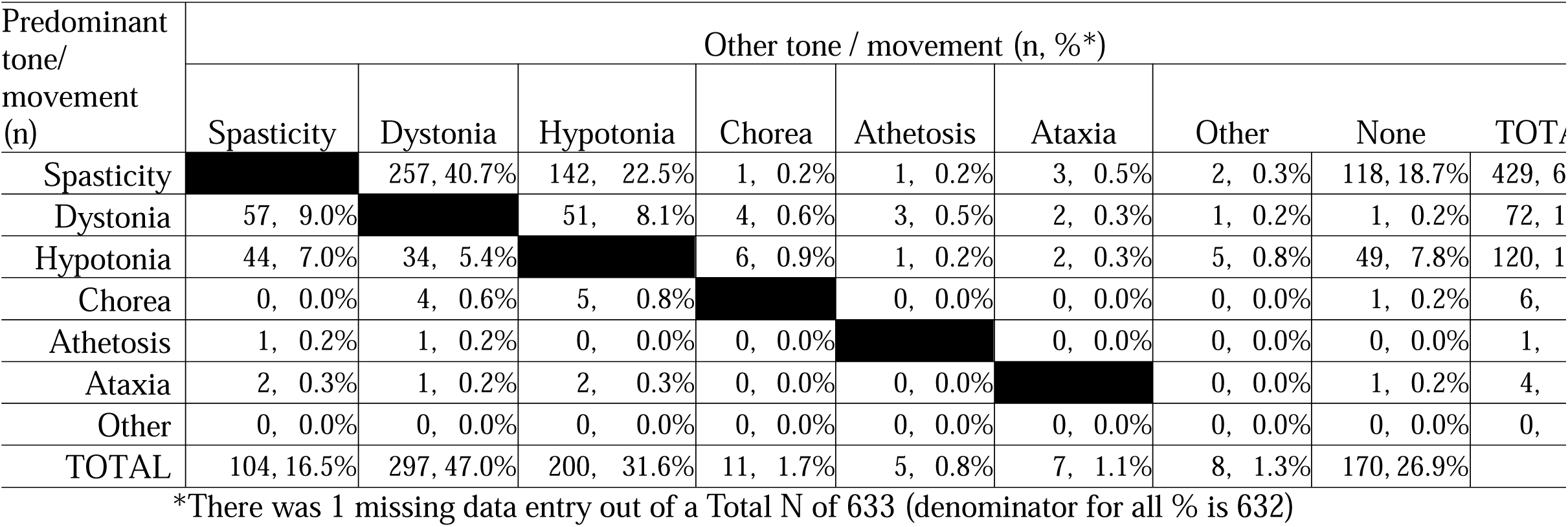
Tone and movement conditions (clinician-entered)

**Table 3.** Distribution of tone and movement conditions (clinician-entered)

|  | n, %<br>(Total N=633) |
| --- | --- |
| Diplegia | 121 , 19.7% |
| Quadriplegia | 288 , 47.0% |
| Right monoplegia | 3 , 0.5% |
| Right hemiplegia | 56 , 9.1% |
| Right triplegia | 28 , 4.6% |
| Left monoplegia | 5 , 0.8% |
| Left hemiplegia | 82 , 13.4% |
| Left triplegia | 30 , 4.9% |
| Missing | 20 , 3.2% |

Missing data percentages are calculated with a denominator of N=633. Other percentages are calculated with a denominator of 633 – n of missing data.

The majority were independently ambulatory (368/633, 58.1%), had minimal manual ability limitations (317/468, 67.7%, at MACS Levels I-II), intact visual function (284/564, 50.4%, at VFCS Level I), and were able to eat all food and drink safely orally (288/515, 55.9%, at EDACS Level I). The majority also had minimal limitations in motor speech output (249/467, 53.3% at VSS levels III and IV) or communication overall (282/539, 52.3% at CFCS levels I and II) (Table 4).

**Table 4.** Functional abilities (clinician-entered)

|  |  |  |
| --- | --- | --- |
| GMFCS (n,%) (Age 0+, N=633) |  |  |
| I | 100 | , 15.8% |
| II | 194 | , 30.6% |
| III | 74 | , 11.7% |
| IV | 117 | , 18.5% |
| V | 148 | , 23.4% |
| Missing | 0 | , 0.0% |
| MACS (n, %) (Age 4+, N=516) |  |  |
| I | 130 | , 27.8% |
| II | 187 | , 40.0% |
| III | 50 | , 10.7% |
| IV | 42 | , 9.0% |
| V | 59 | , 12.6% |
| Missing | 48 | , 9.3% |
| EDACS (n, %) (Age 3+, N=570) |  |  |
| I | 288 | , 55.9% |
| II | 76 | , 14.8% |
| III | 24 | , 4.7% |
| IV | 36 | , 7.0% |
| V | 91 | , 17.7% |
| Missing | 55 | , 9.6% |
| VFCS (n, %) (Age 1+, N=625) |  |  |
| I | 284 | , 50.4% |
| II | 139 | , 24.6% |
| III | 76 | , 13.5% |
| IV | 39 | , 6.9% |
| V | 26 | , 4.6% |
| Missing | 61 | , 9.8% |
| CFCS (n, %) (Age 2+, N=594) |  |  |
| I | 194 | , 36.0% |
| II | 88 | , 16.3% |
| III | 50 | , 9.3% |
| IV | 109 | , 20.2% |
| V | 98 | , 18.2% |
| Missing | 55 | , 9.3% |
| VSS (n, %) (Age 4+, N=516) |  |  |
| I | 139 | , 29.8% |
| II | 110 | , 23.6% |
| III | 75 | , 16.1% |
| IV | 143 | , 30.6% |
| Missing | 49 | , 9.5% |

GMFCS – Gross motor function classification system, MACS – Manual ability classification system, EDACS – Eating and drinking ability classification system, VFCS – Visual function classification system, CFCS – Communication function classification system, VSS – Viking speech scale. Missing data percentages are calculated with a denominator of N=633. Other percentages are calculated with a denominator of 633 – n of missing data.

### Brain MRI patterns, etiology, and birth history

White matter injury was more common (323/618, 52.3%) than grey matter injury (146/618, 23.6%). A minority had cerebral malformations or maldevelopments (92/618, 14.9%) or a normal brain MRI (59/618, 9.5%) (Table 5).

**Table 5.** Brain MRI patterns and etiology (clinician-entered)

|  | n, %<br>(Total N=633) |
| --- | --- |
| MRICS (n) |  |
| Normal | 59 , 9.5% |
| White matter injury - any | 323 , 52.3% |
| White matter injury - PVL | 161 , 26.1% |
| White matter injury - IVH | 96 , 15.5% |
| White matter injury - Other | 111 , 18.0% |
| Grey matter injury - any | 146 , 23.6% |
| Grey matter injury – Basal ganglia/ Thalamus | 43 , 7.0% |
| Grey matter injury – Arterial distribution Infarction | 38 , 6.1% |
| Grey matter injury – Watershed | 7 , 1.1% |
| Grey matter injury - Cortical Other | 79 , 12.8% |
| Malformation/Maldevelopment | 92 , 14.9% |
| MRI images unavailable/not done | 82 , 13.3% |
| Missing | 15 , 2.4% |
| Etiology (n) |  |
| Infection | 34 , 5.4% |
| Prematurity | 251 , 39.7% |
| Stroke | 82 , 13.0% |
| Trauma | 20 , 3.2% |
| Hypoxia-ischemia | 80 , 12.6% |
| Genetic/Metabolic | 146 , 23.1% |
| Unknown | 58 , 9.2% |
| Other | 40 , 6.3% |
| Missing | 0 , 0.0% |

MRICS – MRI Classification System, PVL – periventricular leukomalacia, IVH – intraventricular hemorrhage. Missing data percentages are calculated with a denominator of N=633. Other percentages are calculated with a denominator of 633 – n of missing data.

The single most common CP etiology was prematurity (251/633, 39.7%) followed by genetic/metabolic etiologies (146/633, 23.1%). Of note, 9.2% (58/633) had an unknown etiology, which was typically documented in the absence of neonatal distress (i.e. suggestive of a genetic/metabolic etiology^28,29^) (Table 5).

Over half were born at or after 37 weeks gestation (322/610, 52.8%), vaginally (315/610, 51.6%), and at a birthweight 2500g or greater (296/584, 50.7%). The majority required ICU admission (470/619, 75.9%) (Table 6).

**Table 6.** Birth details (caregiver-entered)

|  | Means, 95% CI or<br>n, % (Total N=633) |
| --- | --- |
| Gestational age, weeks (mean 95% CI) | 34.9 , 34.0 - 35.9 |
| 37 weeks or more (n) | 322 , 52.8% |
| 34-36 weeks (n) | 75 , 12.3% |
| 32-34 weeks (n) | 35 , 5.7% |
| 28-32 weeks (n) | 56 , 9.2% |
| Less than 28 weeks (n) | 122 , 20.0% |
| Missing (n) | 23 , 3.6% |
| Mode of delivery (n) |  |
| Vaginal | 315 , 51.6% |
| Cesarean section | 295 , 48.4% |
| Missing | 23 , 3.6% |
| Birth weight (n) |  |
| Less than 1500 g | 167 , 28.6% |
| 1500-2500 g | 121 , 20.7% |
| 2500-4000 g | 272 , 46.6% |
| More than 4000 g | 24 , 4.1% |
| Missing | 49 , 7.7% |
| Initial ICU stay (n) |  |
| None | 149 , 24.1% |
| Less than 1 month | 179 , 28.9% |
| 1-3 months | 180 , 29.1% |
| 4-6 months | 63 , 10.2% |
| Greater than 6 months | 35 , 5.7% |
| Unspecific duration | 13 , 2.1% |
| Missing | 14 , 2.2% |

Missing data percentages are calculated with a denominator of N=633. Other percentages are calculated with a denominator of 633 – n of missing data.

### Motor medications, botulinum toxin, orthotics, equipment, and therapies

The most common medications people with CP had ever taken for motor symptom management were baclofen (221/633, 34.9%), gabapentin (154/633, 24.3%), diazepam (144/633, 22.7%), and clonidine (121/633, 19.1%). The majority had received botulinum toxin injections (333/633, 52.6%), most commonly in the lower legs (248/623, 39.8%) and upper legs (190/623, 30.5%) (Table 7). The majority wore ankle foot orthoses (458/633 for either left or right, 72.4%) and many used a manual wheelchair (258/633, 40.8%) (Table 8). Most received physical therapy (485/626, 77.5%), occupational therapy (435/626, 69.5%), or speech therapy (369/626, 58.9%), most commonly at school (382/628, 60.8%) via an individualized educational plan (446/627, 71.1%) (Table 9).

**Table 7.** Motor medication and injectables history (caregiver-entered)

|  | n, %<br>(Total N=633) |
| --- | --- |
| Motor medications (n) |  |
| Baclofen | 221 , 34.9% |
| Gabapentin | 154 , 24.3% |
| Diazepam | 144 , 22.7% |
| Clonidine | 121 , 19.1% |
| Levodopa | 55 , 8.7% |
| Trihexyphenidyl | 31 , 4.9% |
| Other | 17 , 2.7% |
| None indicated | 275 , 43.4% |
| Botulinum toxin (n) |  |
| Have had injections | 333 , 52.6% |
| Missing | 0 , 0.0% |
| Injection location |  |
| Upper Arms | 98 , 15.7% |
| Lowers Arms | 97 , 15.6% |
| Hands | 50 , 8.0% |
| Upper Legs | 190 , 30.5% |
| Lower Legs | 248 , 39.8% |
| Feet | 27 , 4.3% |
| Neck | 13 , 2.1% |
| Salivary Glands | 25 , 4.0% |
| Missing | 10 , 1.6% |
| Injection last received |  |
| Less than 3 months ago | 115 , 18.4% |
| 3-6 months ago | 63 , 10.1% |
| 6-12 months ago | 26 , 4.2% |
| More than 1 year ago | 121 , 19.4% |
| Missing | 8 , 1.3% |
| Duration injections have been received |  |
| Less than 1 year | 45 , 7.3% |
| 1-2 years | 47 , 7.6% |
| More than 2 years | 96 , 15.6% |
| Missing | 16 , 7.8% |

Missing data percentages are calculated with a denominator of N=633. Other percentages are calculated with a denominator of 633 – n of missing data.

**Table 8.** Orthotics and equipment (caregiver-entered)

|  | n, %<br>(Total N=633) |
| --- | --- |
| Orthotics (n) |  |
| Left Ankle-foot orthosis | 404 , 63.8% |
| Right Ankle-foot orthosis | 404 , 63.8% |
| Left Supra-malleolar orthosis | 82 , 13.0% |
| Right Supra-malleolar orthosis | 74 , 11.7% |
| Left hand splint | 130 , 20.5% |
| Right hand splint | 125 , 19.7% |
| Left knee immobilizer | 66 , 10.4% |
| Right knee immobilizer | 72 , 11.4% |
| Left stretching split | 90 , 14.2% |
| Right stretching splint | 96 , 15.2% |
| Left shoe insert | 88 , 13.9% |
| Right shoe insert | 93 , 14.7% |
| Trunk support vest | 124 , 19.6% |
| None indicated | 57 , 9.0% |
| Equipment (n) |  |
| Communication Device / Picture Board | 132 , 20.9% |
| Manual Wheelchair | 258 , 40.8% |
| Motorized Wheelchair | 33 , 5.2% |
| Adaptive Stroller | 133 , 21.0% |
| Stander - Less than 5 hours a week | 91 , 14.4% |
| Stander - 5-9 hours a week | 75 , 11.8% |
| Stander - 10 hours or more a week | 26 , 4.1% |
| Stander - usage unspecified | 5 , 0.8% |
| Gait trainer | 117 , 18.5% |
| Walker | 94 , 14.8% |
| Feeding Chair | 65 , 10.3% |
| Bath Chair | 206 , 32.5% |
| Lift (e.g. Hoyer) | 33 , 5.2% |
| None indicated | 173 , 27.3% |

Missing data percentages are calculated with a denominator of N=633. Other percentages are calculated with a denominator of 633 – n of missing data.

**Table 9.**
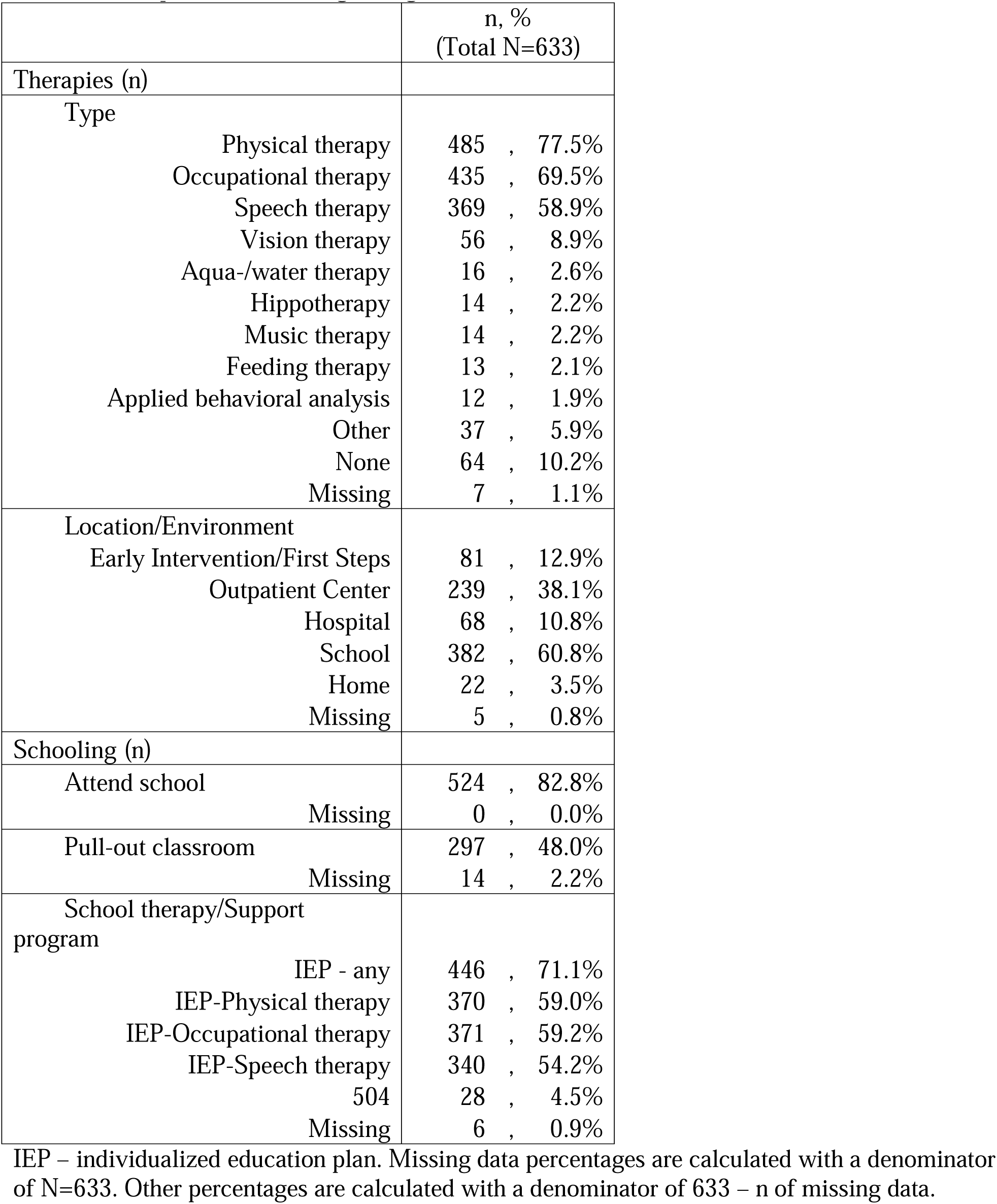
Therapies and schooling (caregiver-entered)

### Medical specialists, surgeries, and nutrition

The most common non-neurologic specialties regularly seen were ophthalmology (273/633, 43.1%), orthopedic surgery (214/633, 33.8%), gastroenterology (182/633, 28.8%), and neurosurgery (142/633, 22.4%). The most common neurosurgical and orthopedic interventions were ventricular shunt placement (102/633, 16.1%) and lower extremity soft tissue surgery (172/633, 27.2%), respectively. The most common general surgery was gastrostomy/gastrojejunostomy tube placement (208/633, 32.9%), with 30.0% (176/586) receiving nutrition via tube feeding (Table 10).

**Table 10.**
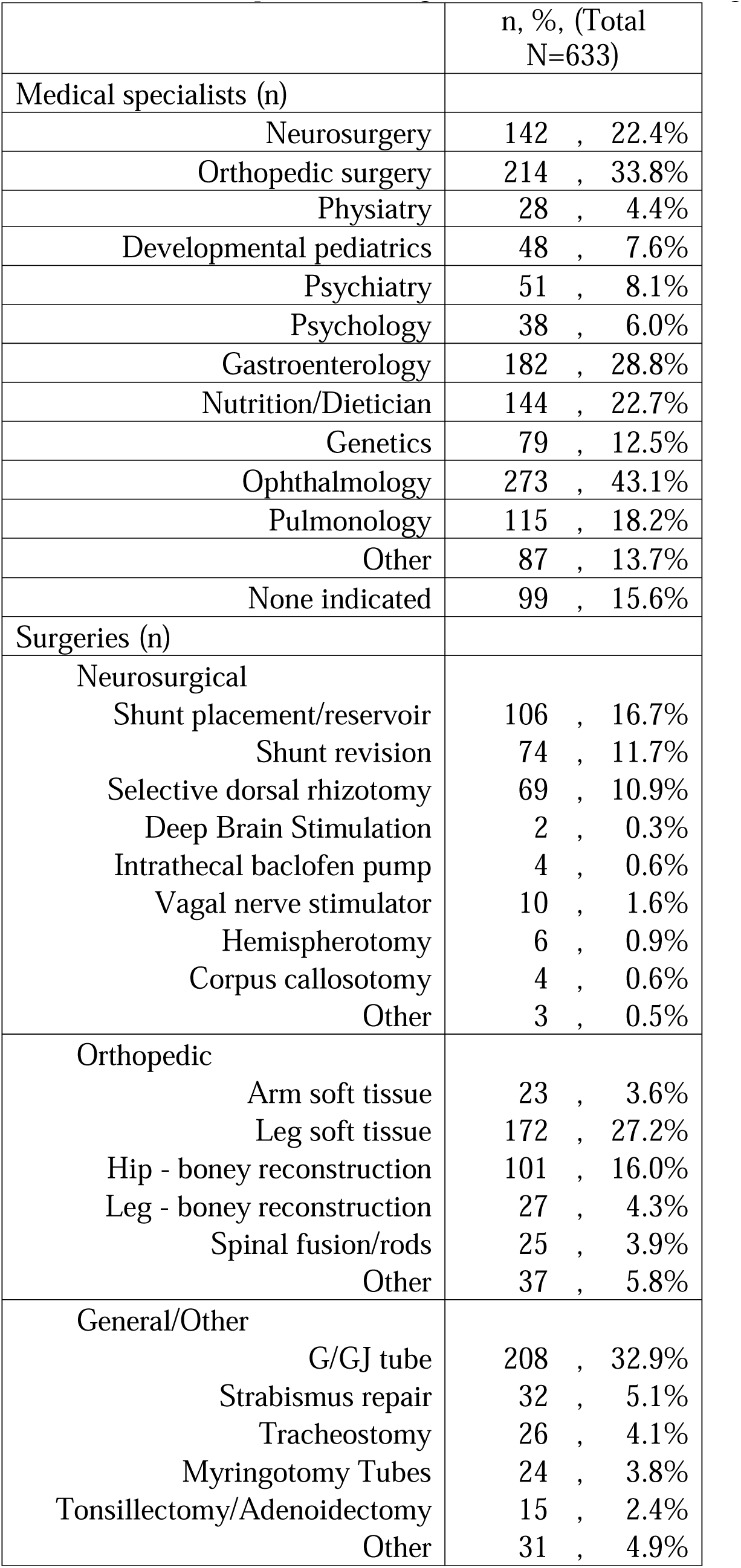
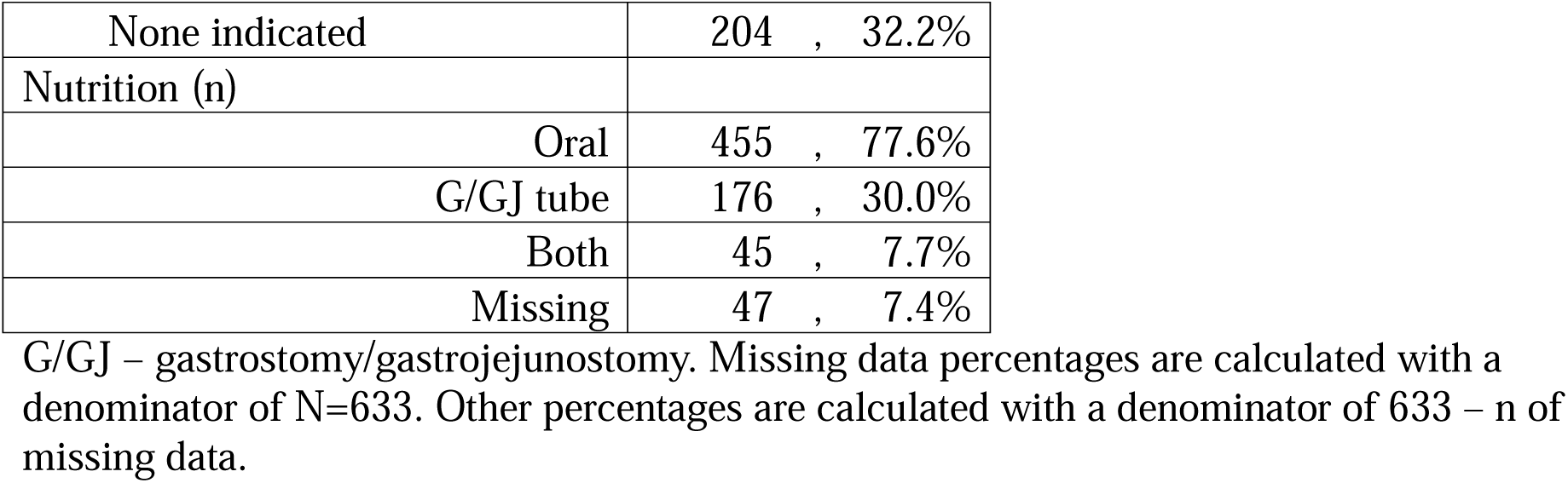
Medical specialists, surgeries, and nutrition (caregiver-entered)

|  | n, %, (Total<br>N=633) |
| --- | --- |
| Medical specialists (n) |  |
| Neurosurgery | 142 , 22.4% |
| Orthopedic surgery | 214 , 33.8% |
| Physiatry | 28 , 4.4% |
| Developmental pediatrics | 48 , 7.6% |
| Psychiatry | 51 , 8.1% |
| Psychology | 38 , 6.0% |
| Gastroenterology | 182 , 28.8% |
| Nutrition/Dietician | 144 , 22.7% |
| Genetics | 79 , 12.5% |
| Ophthalmology | 273 , 43.1% |
| Pulmonology | 115 , 18.2% |
| Other | 87 , 13.7% |
| None indicated | 99 , 15.6% |
| Surgeries (n) |  |
| Neurosurgical |  |
| Shunt placement/reservoir | 106 , 16.7% |
| Shunt revision | 74 , 11.7% |
| Selective dorsal rhizotomy | 69 , 10.9% |
| Deep Brain Stimulation | 2 , 0.3% |
| Intrathecal baclofen pump | 4 , 0.6% |
| Vagal nerve stimulator | 10 , 1.6% |
| Hemispherotomy | 6 , 0.9% |
| Corpus callosotomy | 4 , 0.6% |
| Other | 3 , 0.5% |
| Orthopedic |  |
| Arm soft tissue | 23 , 3.6% |
| Leg soft tissue | 172 , 27.2% |
| Hip - boney reconstruction | 101 , 16.0% |
| Leg - boney reconstruction | 27 , 4.3% |
| Spinal fusion/rods | 25 , 3.9% |
| Other | 37 , 5.8% |
| General/Other |  |
| G/GJ tube | 208 , 32.9% |
| Strabismus repair | 32 , 5.1% |
| Tracheostomy | 26 , 4.1% |
| Myringotomy Tubes | 24 , 3.8% |
| Tonsillectomy/Adenoidectomy | 15 , 2.4% |
| Other | 31 , 4.9% |
| None indicated | 204 , 32.2% |
| Nutrition (n) |  |
| Oral | 455 , 77.6% |
| G/GJ tube | 176 , 30.0% |
| Both | 45 , 7.7% |
| Missing | 47 , 7.4% |

G/GJ – gastrostomy/gastrojejunostomy. Missing data percentages are calculated with a denominator of N=633. Other percentages are calculated with a denominator of 633 – n of missing data.

### Seizure history and other co-existing symptoms

The majority had experienced seizures (327/632, 51.7%). Of people taking anti-seizure medications (268/633, 42.3%), the majority were taking levetiracetam (148/268, 55.2%) followed by clobazam (104/268, 38.8%). As an estimate of rates of drug resistant epilepsy,^30^ 14.4% (83/575) were on at least two anti-seizure medications and had a seizure in the previous 30 days and 8.3% (48/575) were on at least three anti-seizure medications and had a seizure in the previous 30 days. (Table 11).

**Table 11.**
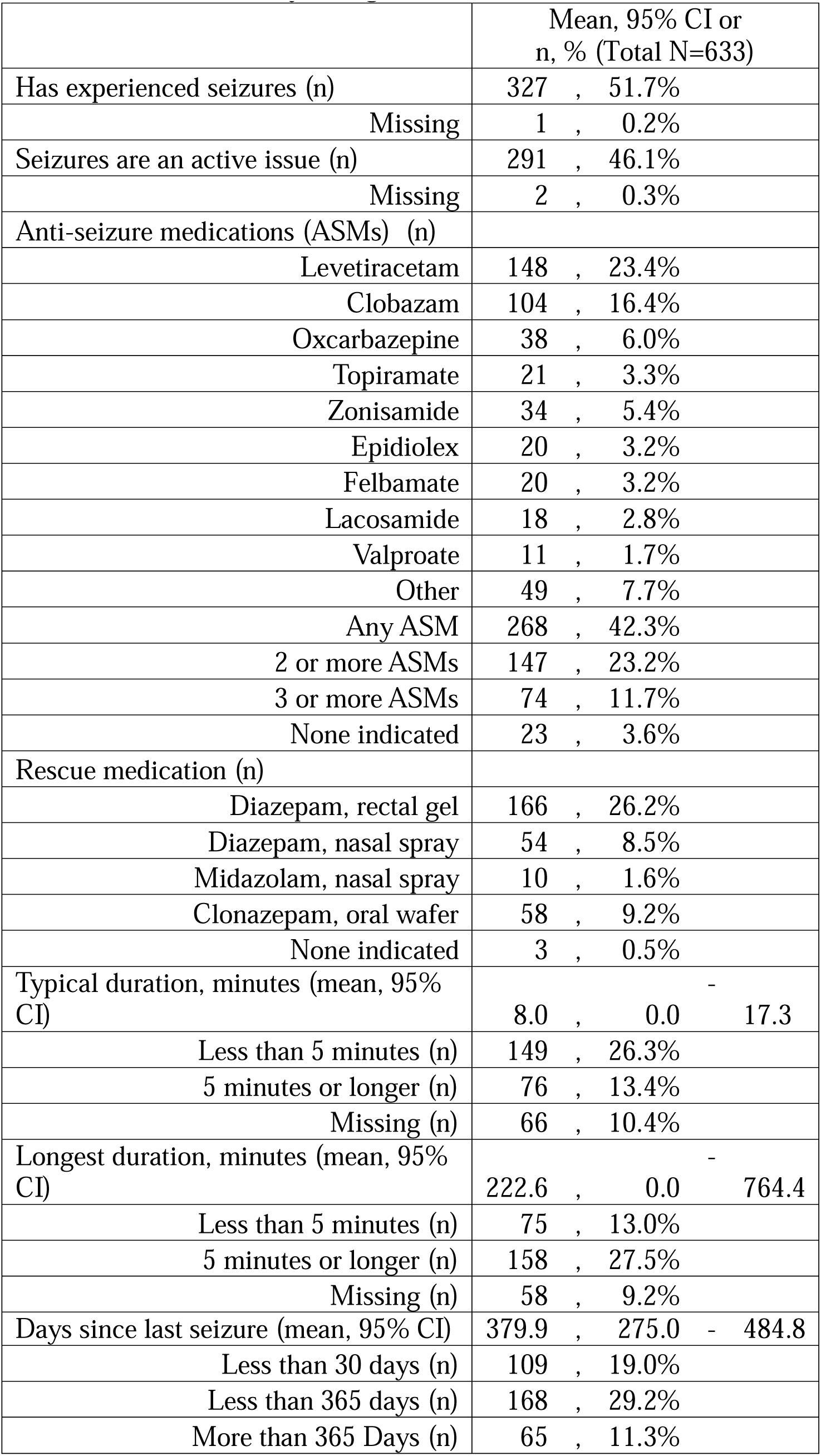
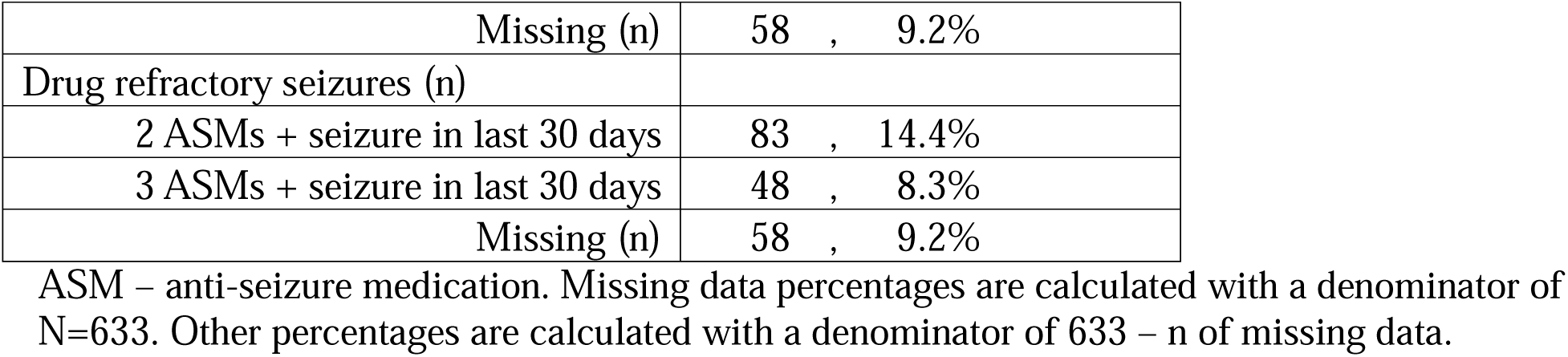
Seizure history (caregiver-entered)

|  | Mean, 95% CI or<br>n, % (Total N=633) |  |  |
| --- | --- | --- | --- |
| Has experienced seizures (n) | 327 | , | 51.7% |
| Missing | 1 | , | 0.2% |
| Seizures are an active issue (n) | 291 | , | 46.1% |
| Missing | 2 | , | 0.3% |
| Anti-seizure medications (ASMs) (n) |  |  |  |
| Levetiracetam | 148 | , | 23.4% |
| Clobazam | 104 | , | 16.4% |
| Oxcarbazepine | 38 | , | 6.0% |
| Topiramate | 21 | , | 3.3% |
| Zonisamide | 34 | , | 5.4% |
| Epidiolex | 20 | , | 3.2% |
| Felbamate | 20 | , | 3.2% |
| Lacosamide | 18 | , | 2.8% |
| Valproate | 11 | , | 1.7% |
| Other | 49 | , | 7.7% |
| Any ASM | 268 | , | 42.3% |
| 2 or more ASMs | 147 | , | 23.2% |
| 3 or more ASMs | 74 | , | 11.7% |
| None indicated | 23 | , | 3.6% |
| Rescue medication (n) |  |  |  |
| Diazepam, rectal gel | 166 | , | 26.2% |
| Diazepam, nasal spray | 54 | , | 8.5% |
| Midazolam, nasal spray | 10 | , | 1.6% |
| Clonazepam, oral wafer | 58 | , | 9.2% |
| None indicated | 3 | , | 0.5% |
| Typical duration, minutes (mean, 95% CI) | 8.0 | , | 0.0 - 17.3 |
| Less than 5 minutes (n) | 149 | , | 26.3% |
| 5 minutes or longer (n) | 76 | , | 13.4% |
| Missing (n) | 66 | , | 10.4% |
| Longest duration, minutes (mean, 95% CI) | 222.6 | , | 0.0 - 764.4 |
| Less than 5 minutes (n) | 75 | , | 13.0% |
| 5 minutes or longer (n) | 158 | , | 27.5% |
| Missing (n) | 58 | , | 9.2% |
| Days since last seizure (mean, 95% CI) | 379.9 | , | 275.0 - 484.8 |
| Less than 30 days (n) | 109 | , | 19.0% |
| Less than 365 days (n) | 168 | , | 29.2% |
| More than 365 Days (n) | 65 | , | 11.3% |

|  |  |
| --- | --- |
| Missing (n) | 58 , 9.2% |
| Drug refractory seizures (n) |  |
| 2 ASMs + seizure in last 30 days | 83 , 14.4% |
| 3 ASMs + seizure in last 30 days | 48 , 8.3% |
| Missing (n) | 58 , 9.2% |

ASM – anti-seizure medication. Missing data percentages are calculated with a denominator of N=633. Other percentages are calculated with a denominator of 633 – n of missing data.

Many expressed pain concerns (230/629, 36.6%), poor sleep (190/629, 30.0%), anxiety (157/633, 24.8%), and attentional difficulty (156/633, 24.6%). Other concerns included avoidance or seeking of loud noises (195/633, 30.8%) and struggling to accept changes in routine (166/633, 26.2%) (Table 12).

**Table 12.**
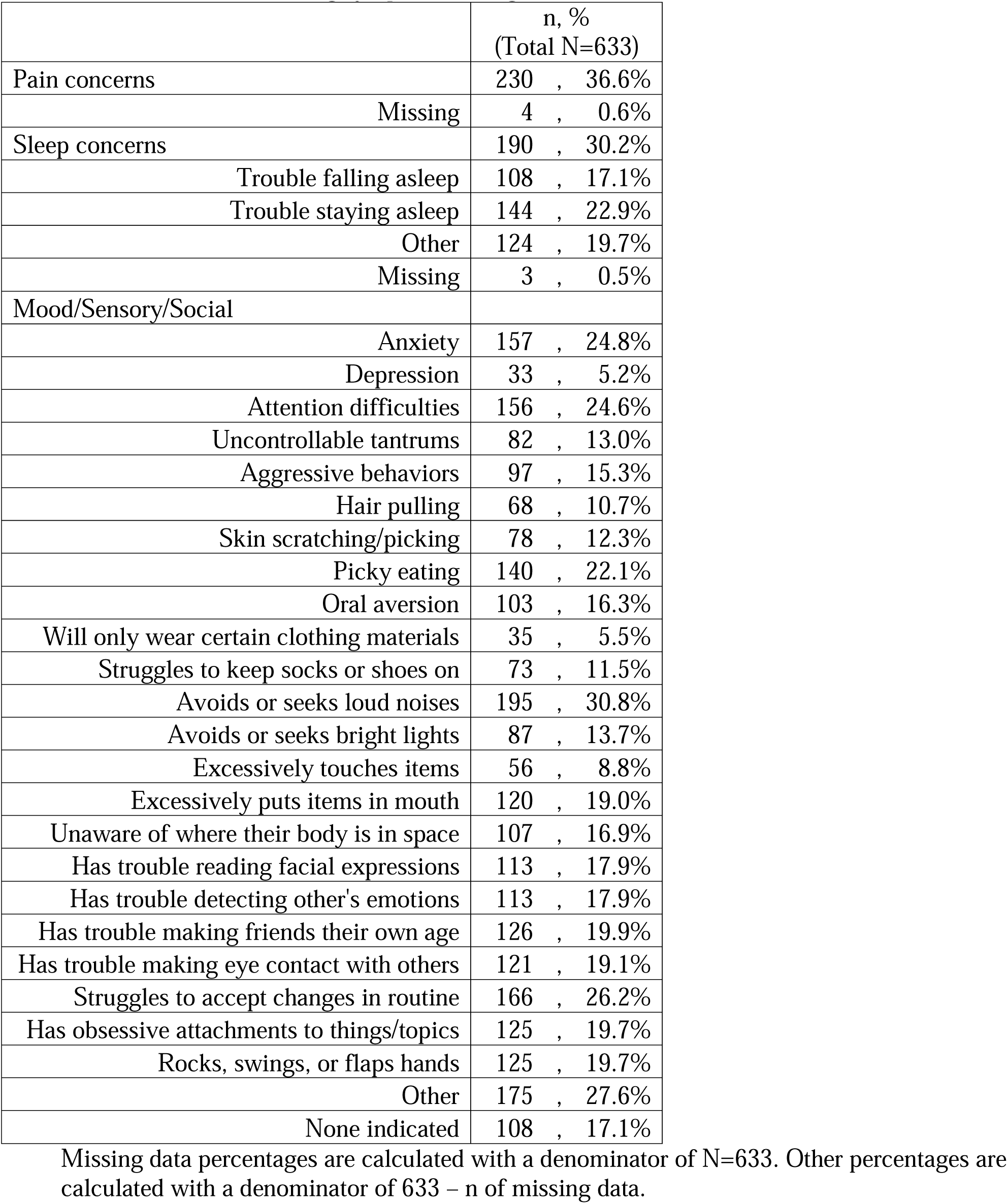
Other co-existing symptoms (caregiver-entered)

|  | n, %<br>(Total N=633) |
| --- | --- |
| Pain concerns | 230 , 36.6% |
| Missing | 4 , 0.6% |
| Sleep concerns | 190 , 30.2% |
| Trouble falling asleep | 108 , 17.1% |
| Trouble staying asleep | 144 , 22.9% |
| Other | 124 , 19.7% |
| Missing | 3 , 0.5% |
| Mood/Sensory/Social |  |
| Anxiety | 157 , 24.8% |
| Depression | 33 , 5.2% |
| Attention difficulties | 156 , 24.6% |
| Uncontrollable tantrums | 82 , 13.0% |
| Aggressive behaviors | 97 , 15.3% |
| Hair pulling | 68 , 10.7% |
| Skin scratching/picking | 78 , 12.3% |
| Picky eating | 140 , 22.1% |
| Oral aversion | 103 , 16.3% |
| Will only wear certain clothing materials | 35 , 5.5% |
| Struggles to keep socks or shoes on | 73 , 11.5% |
| Avoids or seeks loud noises | 195 , 30.8% |
| Avoids or seeks bright lights | 87 , 13.7% |
| Excessively touches items | 56 , 8.8% |
| Excessively puts items in mouth | 120 , 19.0% |
| Unaware of where their body is in space | 107 , 16.9% |
| Has trouble reading facial expressions | 113 , 17.9% |
| Has trouble detecting other's emotions | 113 , 17.9% |
| Has trouble making friends their own age | 126 , 19.9% |
| Has trouble making eye contact with others | 121 , 19.1% |
| Struggles to accept changes in routine | 166 , 26.2% |
| Has obsessive attachments to things/topics | 125 , 19.7% |
| Rocks, swings, or flaps hands | 125 , 19.7% |
| Other | 175 , 27.6% |
| None indicated | 108 , 17.1% |

Missing data percentages are calculated with a denominator of N=633. Other percentages are calculated with a denominator of 633 – n of missing data.

### Family history

A minority had siblings with health concerns (107/610, 17.5%), most commonly attentional difficulties (26/610, 4.3%), autism (19/610, 3.1%), or CP or other motor conditions (18/610, 3.0%). Family history was otherwise notable for autism (70/633, 11.1%), CP (46/633, 7.3%), and seizures/epilepsy (45/633, 7.1%) (Table 13).

**Table 13.** Family history (caregiver-entered)

|  | n, %<br>(Total N=633) |
| --- | --- |
| Siblings | 485 , 78.1% |
| Missing | 12 , 1.9% |
| Siblings with health concerns | 107 , 17.5% |
| Anxiety | 15 , 2.5% |
| Depression | 6 , 1.0% |
| Attention difficulties | 26 , 4.3% |
| Autism | 19 , 3.1% |
| Intellectual disability | 12 , 2.0% |
| CP or motor conditions | 18 , 3.0% |
| Same/similar condition | 8 , 1.3% |
| Genetic condition | 3 , 0.5% |
| Seizures/Epilepsy | 6 , 1.0% |
| Hearing/Vision | 6 , 1.0% |
| Missing | 23 , 3.6% |
| Other family history |  |
| CP | 46 , 7.3% |
| Seizures/Epilepsy | 45 , 7.1% |
| Autism | 70 , 11.1% |
| Intellectual disability | 37 , 5.8% |
| Other childhood-onset condition | 20 , 3.2% |
| None indicated | 486 , 76.8% |

Missing data percentages are calculated with a denominator of N=633. Other percentages are calculated with a denominator of 633 – n of missing data.

### Variables affecting the odds of independent ambulation, oral nutritive feeding, and speech

Binary logistic regression revealed good model fits when assessing whether five variables (ADI, gestational age, birth weight, MRICS brain injury pattern, or etiology) affected the odds of each of three outcomes: independent ambulation (Hosmer-Lemeshow p=0.96, Chi-square p<0.001), the ability to take nutrition orally (Hosmer-Lemeshow p=0.46, Chi-square p<0.001), and the motor ability to produce understandable speech (Hosmer-Lemeshow p=0.75, Chi-square p<0.001) by 5 years old.

Variables that decreased the odds of independent walking were cortical grey matter injury (OR 3.9, 95% CI 1.4-11.3, p=0.01), cerebral malformations (OR 3.5, 95% CI 1.3-9.7, p=0.02), basal ganglia or thalamic injury (OR 7.9, 95% CI 1.7-36.1, p=0.007), infection as a CP etiology (OR 11.6, 95% CI 2.0-66.8, p=0.006), and initial ICU stay between 1-3 months (OR 3.4, 95% CI 1.3-8.7, p=0.01), 4-6 months (OR 6.4, 95% CI 1.8-22.8, p=0.005), or greater than 6 months (OR 8.4, 95% CI 2.1-34.0, p=0.003).

Variables that decreased the odds of oral nutritive intake were cortical grey matter injury (OR 4.1, 95% CI 1.3-13.0, p=0.02), a hypoxia-ischemia CP etiology (OR 4.7, 95% CI 1.3-17.7, p=0.02) and ICU stay between 1-3 months (OR 6.2, 95% CI 1.9-20.5, p=0.003), 4-6 months (OR 17.0, 95% CI 3.2-89.5, p=0.001), or greater than 6 months (OR 32.1, 95% CI 5.6-183.5, p<0.0005).

Variables that decreased the odds of understandable speech production were cortical grey matter injury (OR 8.5, 95% CI 2.7-26.8, p<0.0005), cerebral malformations (OR 6.9, 95% CI 2.3-20.9, p=0.001), intraventricular hemorrhage (OR 4.5, 95% CI 1.6-12.2, p=0.0004), periventricular leukomalacia (OR 3.3, 95% CI 1.4-7.9, p=0.006), an infection etiology (OR 25.6, 95% CI 2.5-259.9, p=0.006), a genetic etiology (OR 7.5, 95% CI 2.2-25.8, p=0.001), or ICU stay greater than 6 months (OR 6.6, 95% CI 1.4-31.1, p=0.02) (Table 14).

**Table 14.** Variables affecting the odds of independent walking, oral nutritive intake, and understandable motor speech by age 5.

| Characteristics that can be determined in infancy | GMFCS IV-V<br>(relative to GMFCS I-III) |  | G/GJ-tube feeding<br>(relative to oral feeding) |  | VSS III-IV<br>(relative to VSS I-II) |  |
| --- | --- | --- | --- | --- | --- | --- |
|  | OR (95% CI) | *p* | OR (95% CI) | *p* | OR (95% CI) | *p* |
| MRICS - Malformation/<br>Maldevelopment | 3.5, 1.3-9.7 | **0.02\*** | 2.9, 0.8-10.0 | 0.09 | 6.9, 2.3-20.9 | **0.001\*** |
| MRICS - Normal | 0.6, 0.2-2.4 | 0.5 | 1.4, 0.3-6.9 | 0.7 | 1.1, 0.3-4.0 | 0.872 |
| MRICS - WM Other | 1.5, 0.6-3.4 | 0.4 | 1.4, 0.5-3.7 | 0.5 | 1.2, 0.5-2.9 | 0.747 |
| MRICS - WM IVH | 1.4, 0.6-3.6 | 0.4 | 0.9, 0.3-3.0 | 0.9 | 4.5, 1.6-12.2 | **0.004\*** |
| MRICS - WM PVL | 2.1, 0.9-4.6 | 0.07 | 1.4, 0.5-3.6 | 0.5 | 3.3, 1.4-7.9 | **0.006\*** |
| MRICS - GM Cortical | 3.9, 1.4-11.3 | **0.01\*** | 4.1, 1.3-13.0 | **0.02\*** | 8.5, 2.7-26.8 | **0.000\*** |
| MRICS - GM Basal Ganglia<br>/ Thalamus | 7.9, 1.7-36.1 | **0.01\*** | 0.6, 0.1-3.2 | 0.6 | 2.6, 0.5-13.0 | 0.239 |
| MRICS - GM AD Infarction | 0.7, 0.1-6.0 | 0.8 | 1.1, 0.1-12.2 | 0.9 | 3.2, 0.5-20.0 | 0.206 |
| MRICS - GM Watershed | 1.5e9, 0.0- ** | 1.0 | 1.0, 0.1-14.6 | 1.0 | 4.8, 0.3-90.9 | 0.294 |
| Etiology - Infection | 11.6, 2.0-66.8 | **0.006\*** | 3.7, 0.9-15.2 | 0.074 | 25. 2.5-259.<br>6, 9 | **0.006\*** |
| Etiology - Prematurity | 0.9, 0.3-3.1 | 0.9 | 0.7, 0.2-2.6 | 0.549 | 0.6, 0.1-2.6 | 0.513 |
| Etiology - Stroke | 0.2, 0.1-1.0 | 0.05 | 1.3, 0.3-5.9 | 0.711 | 0.4, 0.1-1.9 | 0.275 |
| Etiology - Trauma | 1.7, 0.2-13.1 | 0.6 | 3.4, 0.3-35.1 | 0.302 | 12. 0.8-192.<br>6, 4 | 0.068 |
| Etiology - Genetic | 1.7, 0.6-4.8 | 0.3 | 3.1, 1.0-9.9 | 0.050 | 7.5, 2.2-25.8 | **0.001\*** |
| Etiology - Unknown | 1.2, 0.3-4.8 | 0.8 | 3.6, 0.7-18.7 | 0.123 | 1.4, 0.3-6.8 | 0.645 |
| Etiology - HIE | 1.2, 0.4-4.2 | 0.7 | 4.7, 1.3-17.7 | 0.021 | 2.9, 0.7-12.2 | 0.145 |
| Area deprivation index | 1.0, 1.0-1.0 | 0.7 | 1.0, 1.0-1.0 | 0.451 | 1.0, 1.0-1.0 | 0.715 |
| Gestational age (weeks) | 1.0, 0.9-1.1 | 0.7 | 1.0, 0.9-1.2 | 0.723 | 1.1, 0.9-1.2 | 0.245 |
| Delivery (C-Section,<br>relative to vaginal) | 1.1, 0.6-2.0 | 0.7 | 0.5, 0.3-1.1 | 0.103 | 0.9, 0.5-1.7 | 0.835 |
| Birth weight (<1500g,<br>relative to 2500-4000g) | 0.8, 0.2-3.0 | 0.7 | 1.5, 0.2-10.4 | 0.684 | 2.4, 0.5-11.8 | 0.278 |
| Birth weight (1500-2500g,<br>relative to 2500-4000g) | 1.2, 0.5-3.0 | 0.7 | 2.5, 0.8-8.0 | 0.118 | 1.5, 0.5-4.1 | 0.485 |
| Birth weight (>4000g,<br>relative to 2500-4000g) | 0.7, 0.1-3.4 | 0.6 | 0.8, 0.1-5.2 | 0.783 | 0.5, 0.1-2.7 | 0.393 |
| Initial ICU stay (<1 month,<br>relative to none) | 1.6, 0.7-3.9 | 0.3 | 2.0, 0.6-6.2 | 0.233 | 1.5, 0.6-3.6 | 0.377 |
| Initial ICU stay (1-3 months,<br>relative to none) | 3.4, 1.3-8.7 | **0.01\*** | 6.2, 1.9-20.5 | **0.003\*** | 2.3, 0.8-6.2 | 0.108 |
| Initial ICU stay (4-6 months,<br>relative to none) | 6.4, 1.8-22.8 | **0.005\*** | 17.0 3.2-89.5<br>, | **0.001\*** | 3.2, 0.8-12.5 | 0.096 |
| Initial ICU stay (>6 months, | 8.4, 2.1-34.0 | **0.003\*** | 32.1 5.6-183.5 | **0.000\*** | 6.6, 1.4-31.1 | **0.017\*** |
| relative to none) |  |  | , |  |  |  |

\*p<0.05, Wald test, binary logistic regression; \*\*insufficient data to assess. MRICS – MRI Classification system, WM – white matter, GM – grey matter, HIE – hypoxic ischemic encephalopathy, GMFCS – Gross Motor Function Classification System (I-III: independently ambulatory; IV-V: primarily uses a wheelchair to get around), G/GJ – gastrostomy/gastrojejunostomy, VSS – Viking Speech Scale (I-II: motor ability to produce speech that is typically understandable to familiar and unfamiliar listeners; III-IV: limited or no motor ability to produce understandable speech).

## Discussion

We comprehensively captured caregiver and clinician-entered data on 97% of people with CP seen in a tertiary care CP Center and used this data to determine variables affecting the odds of walking, oral feeding, and speech. By sharing our methodology, we aim to facilitate replication of this dataset at other sites (https://bit.ly/CP-Intake-Methodology).

### Racial disparities

Non-White people were likely under-represented in our data. Per US Census data, 43.8%, 75.8%, and 58.3% of residents identify as White in St. Louis City,^31^ Missouri,^32^ and Illinois,^33^ respectively, compared to 83.1% of caregivers and 78.5% of people with CP identifying as White in our data. This is particularly notable given the established higher prevalence of CP in Black children in the US.^1^ Our data highlights our Center’s need to address racial inequities and the ongoing disparities in CP-related data globally.^34,35^

### Comparison to existing datasets

Compared to population-based GMFCS data from Australia^36^, Northern Europe^37^, Canada^38^, and the US^39^ (34-46% Level I, 13-17% Level V) the GMFCS distribution in our data set (15.8% Level I, 23.4% Level V) skewed toward greater gross motor functional limitations. In contrast, the CPRN registry, which primarily aggregates data from tertiary care sites in the US, reports a GMFCS distribution more like ours (20.6% at Level I, 25.5% at Level V).^40^ Therefore, our data set may not be generalizable to population-based estimates, but may be generalizable to how people with CP present at tertiary care centers.

There is less population-based data on other functional classification systems, including none, to our knowledge, from the US. Population-based MACS data from Australia^41^, Northern Europe ^37^, and Canada^42^ (72%-77% at Levels I-III) largely match our MACS distribution (77.7% at Levels I-III).

There has been a growing recognition of mixed motor phenotypes in CP, particularly given variability in CP classification using predominant tone type alone.^43,44^ A single center data set from Australia^45^ showed that 55.2% of people with CP had multiple movement disorders, comparable to our data (59.6%). Descriptions of all tone types present in a person with CP may better describe how CP typically manifests, as opposed to classification using predominant tone type alone.^46^

Rates of epilepsy in people with CP in Australia (37.2%^41^) are comparable to the 42.3% of people in our data set who are on anti-seizure medications. Single center data from Australia suggest that 64% (147/230) of people with CP and epilepsy have drug resistant epilepsy, which differs from our estimate of 31.0% (83/268 people on anti-seizure medications who are on at least 2 anti-seizure medications and had a seizure within the last 30 days). This difference may be due ascertainment bias between the two centers or variable definitions of drug resistant epilepsy (the Australian cohort specified “ongoing seizures”^41^ vs. our specification of a seizure within the last 30 days).

Comparable to recent estimates that 8-30% of people with CP have contributing genetic etiologies,^29,47,48^ 23% of people in our data set had a contributing genetic etiology. Population-based registries vary regarding which genetic conditions should be excluded, regardless of the phenotype.^5,49^ However, given recent affirmations that CP is a phenotypic diagnosis independent of its brain-related etiology,^24–26,48,50,51^ registries may increasingly capture genetic data on people with CP. While this is ongoing, single center data sets can help illuminate genetic contributors to CP^.47,52^

### Variables affecting functional abilities after 5 years old

The relationship between initial ICU stay duration and post-discharge outcomes has been assessed prospectively using ICU cohorts,^53,54^ but with limited data on those with CP or on outcomes after age 2 (limiting assessments of walking, feeding, and speech). In our data, perhaps the best predictor of all assessed functional abilities after age 5 was the duration of the initial ICU stay.

Cortical grey matter injury and basal ganglia and thalamic injury have been implicated in motor developmental outcomes,^55–57^ findings which we re-demonstrated here. There is less data on functional outcomes for people with CP and brain malformations, which we demonstrate affect the odds of independent walking and understandable speech. Etiology also affected the odds of all assessed functional outcomes after age 5, again with little available data in the literature for comparison.

### Future Work

In addition to further analyzing this data set (e.g. differences between people with and without genetic CP etiologies), we should investigate this methodology’s effect on workflow (e.g. length of clinic visits) and medical care (e.g. rates of sleep evaluation and treatment). By sharing our methodology (https://bit.ly/CP-Intake-Methodology), we hope to collaboratively facilitate answering these questions and grow our understanding of how CP manifests.

## Data Availability

De-identified data will be shared with qualified investigators upon request. See https://bit.ly/CP-Intake-Methodology for all materials necessary to replicate our methodology.

## Abbreviations

ADI: area deprivation index
ASM: anti-seizure medication
CFCS: Communication Function Classification System
CP: cerebral palsy
EDACS: Eating and Drinking Ability Classification System
EHR: electronic health record
G/GJ: gastrostomy/gastrojejunostomy
GMFCS: Gross Motor Functional Classification System
IEP: individualized education plan
IVH: intraventricular hemorrhage
MACS: Manual Ability Classification System
MRICS: MRI Classification System
PVL: periventricular leukomalacia
US: United States of America
VFCS: Visual Function Classification System
VSS: Viking Speech Scale.

## References

1. Van Naarden Braun K, Doernberg N, Schieve L, Christensen D, Goodman A, Yeargin-Allsopp M. Birth Prevalence of Cerebral Palsy: A Population-Based Study. Pediatrics. 2016;137(1):1–9. doi:10.1542/peds.2015-2872

2. Hurvitz EA, Gross PH, Gannotti ME, Bailes AF, Horn SD. Registry-based Research in Cerebral Palsy: The Cerebral Palsy Research Network. Phys Med Rehabil Clin N Am. 2020;31(1):185–194. doi:10.1016/j.pmr.2019.09.005

3. Registry Sites | Cerebral Palsy Research Network. Accessed July 15, 2024. https://cprn.org/research/cerebral-palsy-registry-sites/

4. CPRN Document Download | Cerebral Palsy Research Network. Accessed July 15, 2024. https://cprn.org/cprn-document-download/?doc=/data-submission/CPRN-Data-Submission-Specification-v2dot2dot3.pdf&docname=Data%20Submission%20Specification

5. Goldsmith S, Smithers-Sheedy H, Almasri N, et al. Cerebral palsy registers around the world: A survey. Dev Med Child Neurol. 2024;66(6):765–777. doi:10.1111/DMCN.15798

6. Noritz G, Davidson L, Steingass K. Providing a Primary Care Medical Home for Children and Youth With Cerebral Palsy. Pediatrics. 2022;150(6). doi:10.1542/PEDS.2022-060055

7. Sanger TD, Delgado MR, Gaebler-Spira D, Hallett M, Mink JW. Classification and definition of disorders causing hypertonia in childhood. In: Pediatrics. Vol 111. American Academy of Pediatrics; 2003:e89–e97. doi:10.1542/peds.111.1.e89

8. Sanger TD, Chen D, Delgado MR, et al. Definition and Classification of Negative Motor Signs in Childhood. Pediatrics. 2006;118(5):2159–2167. doi:10.1542/peds.2005-3016

9. Sanger TD, Chen D, Fehlings DL, et al. Definition and classification of hyperkinetic movements in childhood. Movement Disorders. 2010;25(11):1538–1549. doi:10.1002/mds.23088

10. Jaleel F, Rust A, Cheung S, et al. Caregiver descriptions of dystonia in cerebral palsy. Ann Clin Transl Neurol. 2024;11(2):242–250. doi:10.1002/ACN3.51941

11. Aravamuthan BR, Ueda K, Miao H, Gilbert L, Smith SE, Pearson TS. Gait features of dystonia in cerebral palsy. Dev Med Child Neurol. 2021;63(6):748–754. doi:10.1111/DMCN.14802

12. Aravamuthan BR, Pearson TS, Ueda K, et al. Determinants of gait dystonia severity in cerebral palsy. Dev Med Child Neurol. 2023;65(7):968–977. doi:10.1111/DMCN.15524

13. Gilbert LA, Gandham S, Ueda K, Chintalapati K, Pearson T, Aravamuthan BR. Upper Extremity Dystonia Features in People With Spastic Cerebral Palsy. Neurol Clin Pract. 2023;13(6). doi:10.1212/CPJ.0000000000200207

14. Jethwa A, Mink J, Macarthur C, Knights S, Fehlings T, Fehlings D. Development of the Hypertonia Assessment Tool (HAT): A discriminative tool for hypertonia in children. Dev Med Child Neurol. 2010;52(5). doi:10.1111/j.1469-8749.2009.03483.x

15. Palisano R, Rosenbaum P, Walter S, Russell D, Wood E, Galuppi B. Development and reliability of a system to classify gross motor function in children with cerebral palsy. Dev Med Child Neurol. 1997;39(4):214–223. doi:10.1111/j.1469-8749.1997.tb07414.x

16. CanChild. Accessed July 29, 2024. https://canchild.ca/en/resources/42-gross-motor-function-classification-system-expanded-revised-gmfcs-e-r

17. Eliasson AC, Krumlinde-Sundholm L, Rösblad B, et al. The Manual Ability Classification System (MACS) for children with cerebral palsy: Scale development and evidence of validity and reliability. Dev Med Child Neurol. 2006;48(7):549–554. doi:10.1017/S0012162206001162

18. Hidecker MJC, Paneth N, Rosenbaum PL, et al. Developing and validating the Communication Function Classification System for individuals with cerebral palsy. Dev Med Child Neurol. 2011;53(8):704–710. doi:10.1111/j.1469-8749.2011.03996.x

19. Pennington L, Virella D, Mjøen T, et al. Development of The Viking Speech Scale to classify the speech of children with cerebral palsy. Res Dev Disabil. 2013;34(10):3202–3210. doi:10.1016/J.RIDD.2013.06.035

20. Baranello G, Signorini S, Tinelli F, et al. Visual Function Classification System for children with cerebral palsy: development and validation. Dev Med Child Neurol. Published online January 1, 2019. doi:10.1111/dmcn.14270

21. Sellers D, Mandy A, Pennington L, Hankins M, Morris C. Development and reliability of a system to classify the eating and drinking ability of people with cerebral palsy. Dev Med Child Neurol. 2014;56(3):245–251. doi:10.1111/dmcn.12352

22. Himmelmann K, Horber V, De La Cruz J, et al. MRI classification system (MRICS) for children with cerebral palsy: development, reliability, and recommendations. Dev Med Child Neurol. 2017;59(1):57–64. doi:10.1111/dmcn.13166

23. NIH Common Data Elements (CDE) Repository. Accessed July 18, 2024. https://cde.nlm.nih.gov/home

24. Aravamuthan BR, Fehlings D, Shetty S, et al. Variability in Cerebral Palsy Diagnosis. Pediatrics. 2021;147(2):e2020010066. doi:10.1542/peds.2020-010066

25. Rosenbaum P, Paneth N, Leviton A, et al. A report: the definition and classification of cerebral palsy April 2006. Dev Med Child Neurol Suppl. 2007;109:8–14. Accessed March 26, 2019. http://www.ncbi.nlm.nih.gov/pubmed/17370477

26. Aravamuthan BR, Fehlings DL, Novak I, et al. Uncertainties regarding cerebral palsy diagnosis: opportunities to operationalize the consensus definition. Neurol Clin Pract. 2023;in press. doi:10.1101/2023.06.29.23292028

27. Kind AJH, Buckingham WR. Making Neighborhood-Disadvantage Metrics Accessible — The Neighborhood Atlas. New England Journal of Medicine. 2018;378(26):2456–2458. doi:10.1056/NEJMP1802313

28. Aravamuthan BR, Shusterman M, Green Snyder L, Lemmon ME, Bain JM, Gross P. Diagnostic preferences include discussion of etiology for adults with cerebral palsy and their caregivers. Dev Med Child Neurol. 2022;64(6):723–733. doi:10.1111/DMCN.15164

29. Srivastava S, Lewis SA, Cohen JS, et al. Molecular Diagnostic Yield of Exome Sequencing and Chromosomal Microarray in Cerebral Palsy: A Systematic Review and Meta-analysis. JAMA Neurol. 2022;79(12):1287–1295. doi:10.1001/JAMANEUROL.2022.3549

30. Kwan P, Arzimanoglou A, Berg AT, et al. Definition of drug resistant epilepsy: consensus proposal by the ad hoc Task Force of the ILAE Commission on Therapeutic Strategies. Epilepsia. 2010;51(6):1069–1077. doi:10.1111/J.1528-1167.2009.02397.X

31. Census Data | City of St. Louis. Accessed July 25, 2024. https://www.stlouis-mo.gov/government/departments/planning/research/census/data/index.cfm

32. Missouri Demographics - Map of Population by Race - Census Dots. Accessed July 25, 2024. https://www.censusdots.com/race/missouri-demographics

33. Illinois Demographics - Map of Population by Race - Census Dots. Accessed July 25, 2024. https://www.censusdots.com/race/illinois-demographics

34. McIntyre S, Goldsmith S, Webb A, et al. Global prevalence of cerebral palsy: A systematic analysis. Dev Med Child Neurol. Published online August 11, 2022. doi:10.1111/DMCN.15346

35. Avila-Soto F, Kakooza AM, Ueda K, Aravamuthan B. Studies of CP Prevalence: Disparities in Authorship, Citations, and Geographic Location. Pediatr Neurol. 2023;143:59–63. doi:10.1016/J.PEDIATRNEUROL.2023.02.003

36. Reid SM, Carlin JB, Reddihough DS. Using the Gross Motor Function Classification System to describe patterns of motor severity in cerebral palsy. Dev Med Child Neurol. 2011;53(11):1007–1012. doi:10.1111/J.1469-8749.2011.04044.X

37. Hollung SJ, Hägglund G, Gaston MS, et al. Point prevalence and motor function of children and adolescents with cerebral palsy in Scandinavia and Scotland: a CP-North study. Dev Med Child Neurol. 2021;63(6):721. doi:10.1111/DMCN.14764

38. Huroy M, Behlim T, Andersen J, et al. Stability of the Gross Motor Function Classification System over time in children with cerebral palsy. Dev Med Child Neurol. 2022;64(12):1487–1493. doi:10.1111/DMCN.15375

39. Kirby RS, Wingate MS, Van Naarden Braun K, et al. Prevalence and functioning of children with cerebral palsy in four areas of the United States in 2006: a report from the Autism and Developmental Disabilities Monitoring Network. Res Dev Disabil. 2011;32(2):462–469. doi:10.1016/J.RIDD.2010.12.042

40. CPRN DCC Portal | Login. Accessed July 30, 2024. https://my.cprn.org

41. Feroze N, Karim T, Ostojic K, et al. Clinical features associated with epilepsy occurrence, resolution, and drug resistance in children with cerebral palsy: A population-based study. Dev Med Child Neurol. 2024;66(6):793–803. doi:10.1111/DMCN.15807

42. Springer A, Dyck Holzinger S, Andersen J, et al. Profile of children with cerebral palsy spectrum disorder and a normal MRI study. Neurology. 2019;93(1):E88–E96. doi:10.1212/WNL.0000000000007726/ASSET/6B13CCA1-E6DD-4635-94D9-352A84BC8DFD/ASSETS/GRAPHIC/14TTU1A.JPEG

43. Gainsborough M, Surman G, Maestri G, Colver A, Cans C. Validity and reliability of the guidelines of the surveillance of cerebral palsy in Europe for the classification of cerebral palsy. Dev Med Child Neurol. 2008;50(11):828–831. doi:10.1111/J.1469-8749.2008.03141.X

44. Reid SM, Carlin JB, Reddihough DS. Distribution of motor types in cerebral palsy: how do registry data compare? Dev Med Child Neurol. 2011;53(3):233–238. doi:10.1111/J.1469-8749.2010.03844.X

45. Dar H, Stewart K, McIntyre S, Paget S. Multiple motor disorders in cerebral palsy. Dev Med Child Neurol. 2024;66(3):317–325. doi:10.1111/DMCN.15730

46. Christine C, Dolk H, Platt MJ, Colver A, Prasauskiene A, Krageloh-Mann I. Recommendations from the SCPE collaborative group for defining and classifying cerebral palsy. Dev Med Child Neurol Suppl. 2007;109:35–38.

47. May HJ, Fasheun JA, Bain JM, et al. Genetic testing in individuals with cerebral palsy. Dev Med Child Neurol. Published online June 2021. doi:10.1111/dmcn.14948

48. Fahey MC, Maclennan AH, Kretzschmar D, Gecz J, Kruer MC. The genetic basis of cerebral palsy. Dev Med Child Neurol. 2017;59(5):462–469. doi:10.1111/dmcn.13363

49. Smithers-Sheedy H, Badawi N, Blair E, et al. What constitutes cerebral palsy in the twenty-first century? Dev Med Child Neurol. 2014;56(4):323–328. doi:10.1111/dmcn.12262

50. Aravamuthan BR, Shevell M, Kim YM, et al. Role of child neurologists and neurodevelopmentalists in the diagnosis of cerebral palsy: A survey study. Neurology. 2020;95(21):962–972. doi:10.1212/WNL.0000000000011036

51. Aravamuthan BR, Shusterman M, Snyder LG, Lemmon ME, Bain JM, Gross PH. Diagnostic preferences amongst cerebral palsy community members: a cerebral palsy diagnosis should be accompanied by description of its etiology. Dev Med Child Neurol. Published online 2022.

52. Chopra M, Gable DL, Love-Nichols J, et al. Mendelian etiologies identified with whole exome sequencing in cerebral palsy. Ann Clin Transl Neurol. 2022;9(2):193–205. doi:10.1002/acn3.51506

53. Royer ASM, Busari JO. A systematic review of the impact of intensive care admissions on post discharge cognition in children. Eur J Pediatr. 2021;180(12):3443–3454. doi:10.1007/S00431-021-04145-5

54. Subedi D, Deboer MD, Scharf RJ. Developmental trajectories in children with prolonged NICU stays. Arch Dis Child. 2017;102(1):29–34. doi:10.1136/ARCHDISCHILD-2016-310777

55. Martinez-Biarge M, Diez-Sebastian J, Kapellou O, et al. Predicting motor outcome and death in term hypoxic-ischemic encephalopathy. Neurology. 2011;76(24):2055–2061. doi:10.1212/WNL.0b013e31821f442d

56. Kidokoro H, Anderson PJ, Doyle LW, Woodward LJ, Neil JJ, Inder TE. Brain injury and altered brain growth in preterm infants: predictors and prognosis. Pediatrics. 2014;134(2). doi:10.1542/PEDS.2013-2336

57. Anderson PJ, Cheong JLY, Thompson DK. The predictive validity of neonatal MRI for neurodevelopmental outcome in very preterm children. Semin Perinatol. 2015;39(2):147–158. doi:10.1053/J.SEMPERI.2015.01.008

